# SARS-CoV-2 RNA load in the lower respiratory tract, viral RNAemia and N-antigenemia in critically ill adult COVID-19 patients: relationship with biomarkers of disease severity

**DOI:** 10.1101/2021.04.16.21255601

**Authors:** Beatriz Olea, Eliseo Albert, Ignacio Torres, Roberto Gozalvo-Rovira, Nieves Carbonell, José Ferreres, Sandrine Poujois, Rosa Costa, Javier Colomina, Jesús Rodríguez, María Luisa Blasco, David Navarro

## Abstract

**Background:** Little is known about the comparative kinetics of SARS-CoV-RNA load in the lower respiratory tract and in blood compartment in patients admitted to the intensive care unit, and how these relate to biomarkers of COVID-19 severity.

**Methods:** Seventy-three consecutive critically ill COVID-19 patients (median age, 65 years) were recruited. Serial lower respiratory tract (n=165) and plasma (n=340) specimens were collected. RT-PCR and lateral flow immunochromatography assay were used for SARS-CoV-2 RNA quantitation and N protein detection in plasma, respectively. Serum levels of inflammatory and tissue-damage biomarkers in paired specimens were analyzed.

**Results:** SARS-CoV-RNA was detected in the lower respiratory tract of most patients (92%). Viral RNAemia and N-antigenemia were documented in 35.6% and 40.1% of patients, respectively. Viral RNAemia and N-antigenemia cleared at a faster rate than SARS-CoV-2 RNA in tracheal aspirates (TA). SARS-CoV-2 RNA load was higher (*P*<0.001) in TA than in plasma, and correlated significantly (Rho, 0.41; *P*<0.001). A modest correlation was found between SARS-CoV-2 RNA load in TA and plasma and levels of ferritin and lactose dehydrogenase (Rho≤0.3; *P*≤0.008) in paired serum specimens. Neither the dynamics of SARS-CoV-2 RNA load in TA and plasma, nor N-antigenemia detection rate differed between surviving and deceased patients. Yet, a trend towards a higher mortality was seen in patients with viral RNAemia (OR; 2.82; 95% CI, 0.94-8.47; *P*=0.06).

**Conclusion:** Neither SARS-CoV-2 replication rate in the lower respiratory tract nor its presence in the blood appeared to critically impact on survival in ICU COVID-19 patients.

**SUMMARY:** SARS-CoV-2 RNA load in the lower respiratory tract and plasma and N-antigenemia followed different kinetics, correlated modestly with serum levels of inflammatory and tissue-damage biomarkers and lymphopenia and did not appear to increase overall mortality risk in critically ill adult COVID-19 patients.

## INTRODUCTION

Severe COVID-19 is a multisystem disease involving the lower respiratory tract (LRT) and extra-pulmonary organs such as the liver, kidney, spleen and central nervous system [1]. Following infection, SARS-CoV-2 initially replicates in the upper respiratory tract (URT) before reaching the LRT [2,3], where it may cause severe damage, by virtue of its own cytopathogenicity and notably by inducing a persistent, dysregulated pro-inflammatory state [4,5]. While evidence has accumulated suggesting a direct relationship between SARS-CoV-2 RNA load in the URT early after infection and progression to severe clinical forms of COVID-19 [6-8], data linking the extent of virus replication in the LRT with clinical outcomes are scarce [9-11]. In this context, it remains to be elucidated whether acute respiratory distress syndrome and multiple organ failure, frequently ensuing in critically ill patients, are driven by uncontrolled viral replication in the LRT, dysregulated host-immune response, or both. SARS-CoV-2 may access the systemic compartment early after infection, as has been shown for SARS-CoV and MERS [12,13]. In fact, depending upon clinical severity, SARS-CoV-2 RNAemia can be detected in up to 88% of COVID-19 patients within the first week after symptoms onset [14-25], and has been associated with ICU admission, need for invasive mechanical ventilation, multiple organ failure and mortality rate [see meta-analysis in 26]. Likewise, SARS-CoV-2 nucleocapsid (N) antigenemia, which has also been found in a large percentage of COVID-19 patients [27,28], has been associated with higher ICU admission rates and overall mortality [27,29]. To our knowledge, no comprehensive analysis has been published concerning the interplay between the kinetics of SARS-CoV-2 RNA load in the LRT, viral RNAemia and N-antigenemia, and how these relate to the inflammatory state of patients with severe COVID-19. Studies of this nature may contribute in clarifying the pathogenesis of SARS-CoV-2 infection, as well as precisely identifying virological factors modulating COVID-19 prognosis. Here, we addressed these issues in a single-centre cohort of critically ill adult COVID-19 patients.

## PATIENTS AND METHODS

### Patients and specimens

In this prospective observational study, 73 consecutive critically ill COVID-19 patients (51 males and 22 females; median age, 65 years; range, 21 to 80 years) were recruited during ICU stay between October 2020 and February 2021 (Table 1). Respiratory and plasma specimens were scheduled to be collected at least once a week from admission. A total of 195 respiratory specimens (30 nasopharyngeal and 165 tracheal aspirates) and 340 plasma specimens were available for the analyses described below. Medical history and laboratory data were prospectively recorded. The current study was approved by the Ethics Committee of Hospital Clínico Universitario INCLIVA (May,2020).

**Table 1.** Baseline clinical characteristics of the study population at Intensive Care Unit admission.

|  |  |
| --- | --- |
| **Variable** | **no. (%)** |
| **Gender** |  |
| Male | 51 (69.9) |
| Female | 22 (30.1) |
| **Acute physiology and chronic health evaluation (APACHE) II score** |  |
| <10 | 14 (19.2) |
| 10-14 | 27 (37.0) |
| 15-29 | 32 (43.8) |
| **Comorbidities** |  |
| Diabetes mellitus | 18 (24.7) |
| Asthma/ Chronic lung disease | 11 (15.0) |
| Hypertension | 33 (45.2) |
| Obesity | 38 (52.0) |
| Chronic heart disease | 9 (12.3) |
| Vascular disease | 7 (9.6) |
| Cancer | 3 (4.1) |
| Hematologic disease | 3 (4.1) |
| **Number of comorbidity conditions** |  |
| One | 22 (30.1) |
| Two or more | 33 (45.2) |
| None | 18 (24.7) |
| **Oxygenation and ventilator support** |  |
| Invasive mechanical ventilation | 64 (87.7) |
| PiO<sub>2</sub>/FiO<sub>2</sub> <150 mmHg | 58 (79.5) |
| **Acute kidney disfunction** | 17 (23.3) |
| **Antiviral or anti-inflammatory treatment** |  |
| Remdesivir | 16 (21.9) |
| Corticosteroids | 70 (95.9) |
| Tocilizumab | 27 (36.9) |

### Detection of SARS-CoV-2 RNA by RT-PCR

Nasopharyngeal (NP) specimens were placed in 3 ml of Universal Transport Medium (UTM, Becton Dickinson, Sparks, MD, USA). Tracheal aspirates (TA) from mechanically ventilated patients were collected undiluted in sterile containers. Specimens were kept at 4 °C until processed (within 6 hours of receipt). Plasma specimens were obtained by centrifugation of whole blood EDTA tubes, cryopreserved at −80 °C and retrieved for analyses within one month after collection. Non-previously thawed specimens were used for analyses. Nucleic acid extraction was performed using a magnetic microparticle-based protocol (Abbott mSample Preparation SystemDNA) on the Abbott m2000sp platform (Abbott Molecular (Des Plaines, IL, USA) with a starting sample volume of 400 µl. SARS-CoV-2 RNA amplification was carried out by the Abbott Real*Ti*me SARS-CoV-2 assay, a dual target reverse transcription polymerase chain reaction (RT-PCR) assay amplifying the RdRp and N-genes, on the m2000rt platform, following the manufacturer’s instructions. The analytical sensitivity of the RT-PCR assay in TA and plasma specimens was assessed by spiking (in quintuplicate) 10-10^4^ copies/ml of the AMPLIRUN® TOTAL SARS-CoV-2 RNA Control (Vircell SA, Granada, Spain) into RT-PCR negative TA or plasma samples. The limit of detection (LOD) was found to be approximately 100 copies/ml (95%) for both matrices (not shown), in agreement with previous estimates using NP [30,31]. SARS-CoV-2 viral loads are given in copies/ml throughout the study, as estimated using the abovementioned reference material. All specimens from a given patient were analysed in the same run in singlets.

### Detection of SARS-CoV-2 N protein in plasma

The CLINITEST^®^ Rapid COVID-19 Antigen Test (Siemens, Healthineers, Erlangen, Germany), a lateral flow immunochromatography (LFIC) device licensed for detection of SARS-CoV-2 nucleocapsid protein NP or nasal swabs, was used on plasma specimens. The limit of detection (LOD) of the assay in plasma was determined by spiking a pre-pandemic plasma pool testing negative by RT-PCR with 10, 25, 50, 100 and 150 pg/ml of a recombinant N protein (MT-25C19NC, Certest Biotec S.L., Zaragoza, Spain). The LOD was found to be at least 50 pg/ml (Supplementary Fig. 1). Depleting experiments using a rabbit anti-N protein antibody (40143-R019; SinoBiological) confirmed the true nature of SARS-CoV-2 N detected in a number of discordant plasma specimens (testing negative by RT-PCR), as described in supplementary methods and shown in Supplementary Fig. 2.

### Laboratory measurements

Clinical laboratory tests included serum levels of ferritin, D-Dimer (D-D), C reactive protein (CRP), interleukin-6 (IL-6), lactose dehydrogenase (LDH) and absolute lymphocyte counts.

### Statistical methods

Frequency comparisons for categorical variables were carried out using Fisher’s exact test. Differences between medians were compared using the Mann-Whitney U test. Spearman’s rank test was used for analysis of correlation between continuous variables. Two-sided exact *P* values were reported. A *P* value <0.05 was considered statistically significant. The level of agreement between qualitative results provided by paired virological assays was assessed using Kappa-Cohen statistics. Logistic regression analyses were performed to assess risk factors for all-cause mortality. Parameters yielding *P* values <0.1 in univariate analyses were introduced in multivariate models. The analyses were performed using SPSS version 20.0 (SPSS, Chicago, IL, USA).

## RESULTS

### Patient clinical features

Patients were admitted to ICU at a median of 9 days (range, 2–25) after onset of symptoms. All patients presented with pneumonia and imaging findings compatible with COVID-19 on chest-x ray or CT-scan, and most eventually needed mechanical ventilation (87.7%). Median time of ICU stay was 18 days (range, 2–67). A total of 16 and 27 patients were under remdesivir and tocilizumab treatment, respectively, at ICU admission. Corticosteroids were given to 70 patients. A total of 29 patients died, at a median of 22 days (range, 7–66) after ICU admission.

### Kinetics of SARS-CoV-2 RNA load in the lower respiratory tract

Of the 64 patients undergoing mechanical ventilation, 61 had one or more TA collected (total number, 165; median of 2 specimens/patient; range, 1–11). The remaining three patients were not sampled. SARS-CoV-2 RNA was detected in 109 TA from 56 patients (91.8%). The remaining five patients tested negative for SARS-CoV-2 RNA in TA but had one or more positive NP collected prior to mechanical ventilation.

SARS-CoV-2 RNA load in TA ranged between 3.03 and 10.6 log_10_ copies/ml (median, 6.5 log_10_ copies/ml). The dynamics of SARS-CoV-2 RNA load over time in TA is shown in Fig. 1A. Viral load remained relatively stable across the first two weeks from symptoms onset and began to decrease afterwards. No patient tested positive for SARS-CoV-2 RNA in TA beyond day 42.

**Figure 1.**
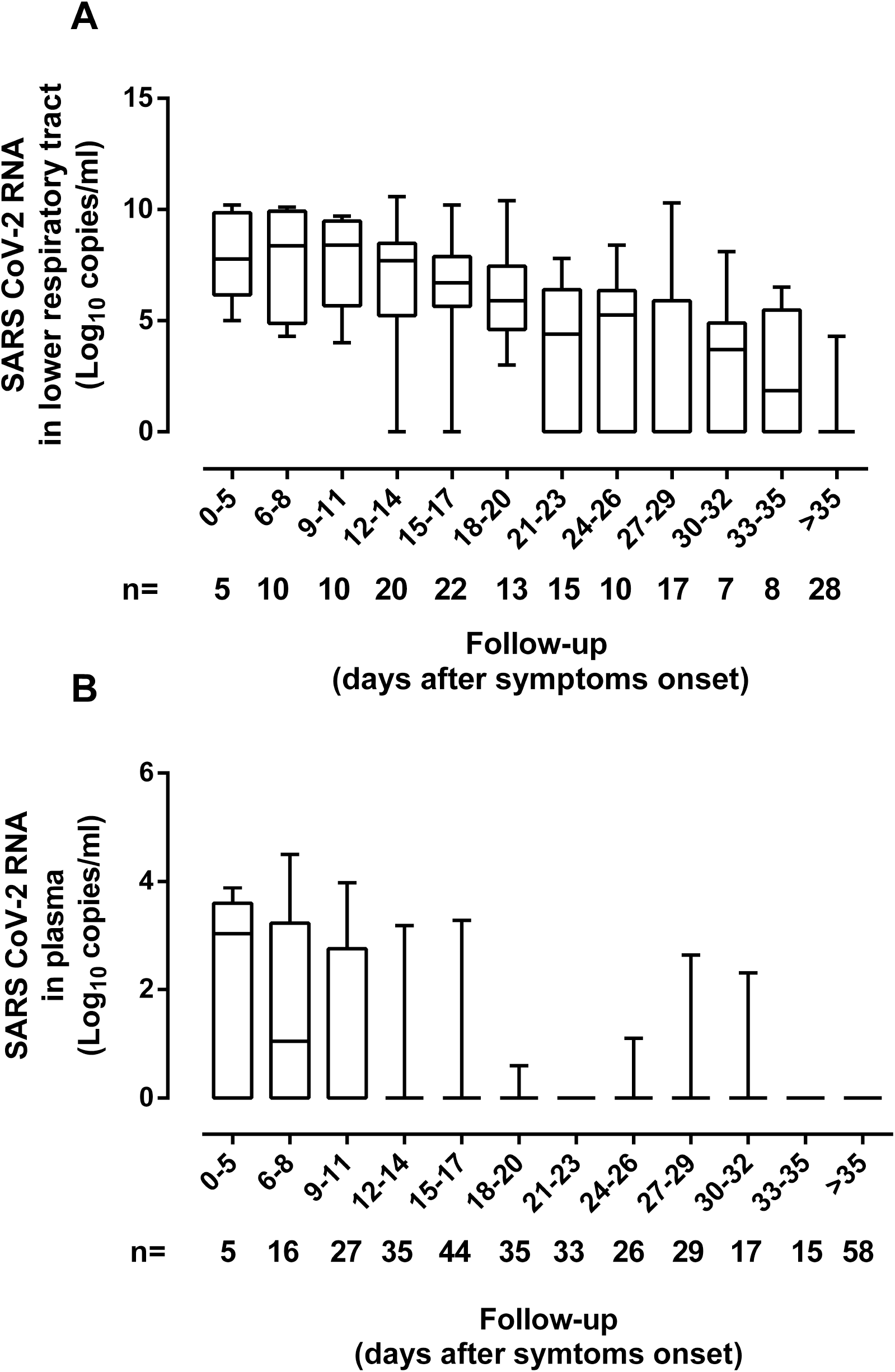
Kinetics of SARS-CoV-RNA load in the lower respiratory tract (tracheal aspirates) (A) and plasma (B) of critically ill patients undergoing invasive ventilation.

Neither remdesivir nor tocilizumab administration appeared to have a major impact on the dynamics of SARS-CoV-2 load in TA (Fig. 2, Supplementary Table 1).

**Figure 2.**
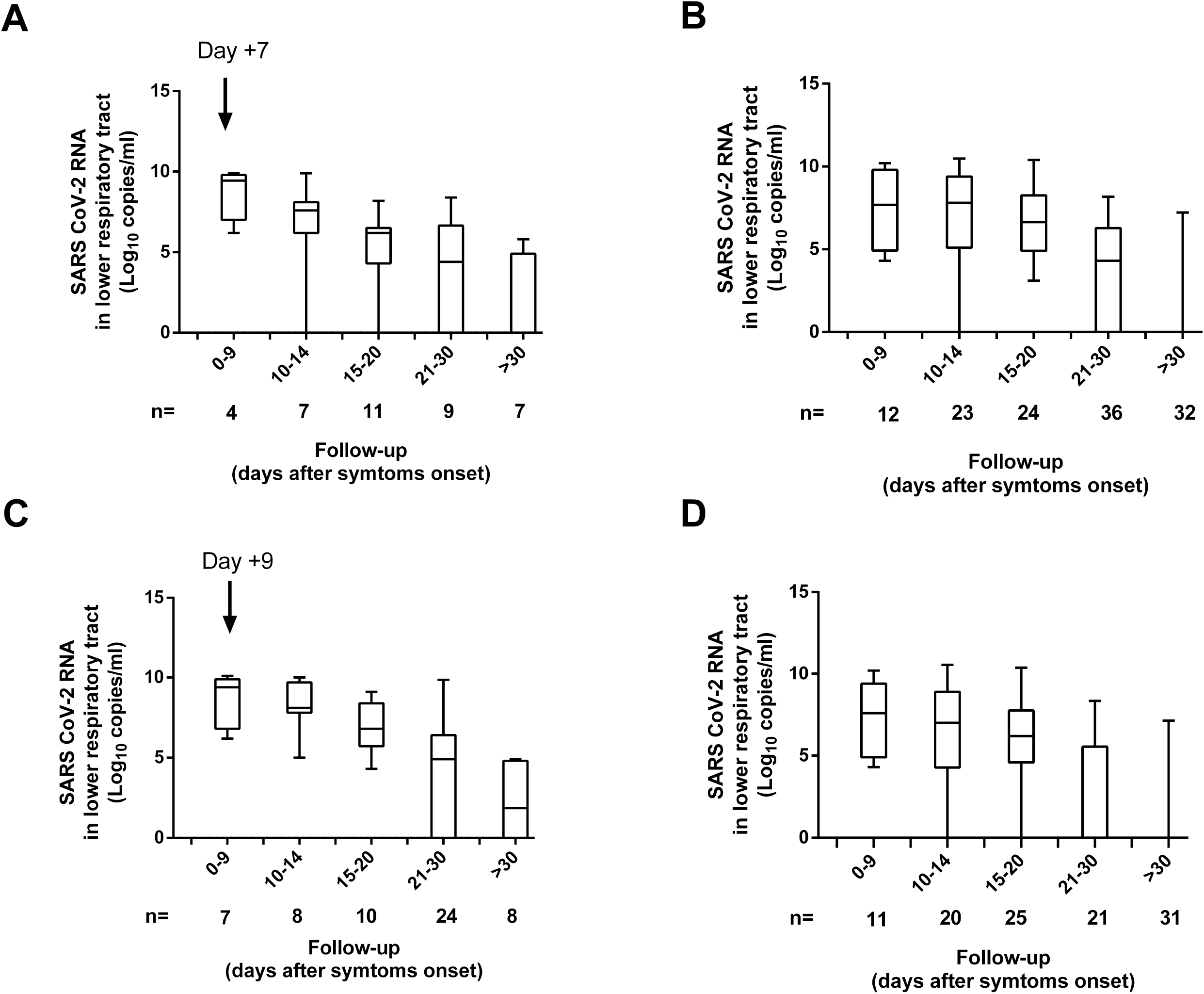
Effect of remdesivir or tocilizumab treatment on the kinetics of SARS-CoV-2 RNA load in the lower respiratory tract (tracheal aspirates) in critically ill patients undergoing invasive ventilation. Panels A and C depict the kinetics of SARS-CoV-2 RNA load in patients receiving remdesivir (A) or tocilizumab (C). The arrow indicates the time of treatment initiation. Remdesivir was administered to 16 patients at a median of 7 days (range, 2–13) since symptoms onset, for a median of 4 days (range, 1–5). Tocilizumab was prescribed to 27 patients (a single dose in 24 patients and two doses in the remaining three patients) at a median of 9 days (range, 3-28) since the onset of symptoms. Panels B and D depict SARS-CoV-2 RNA load kinetics in patients not treated with remdesivir (B) or tocilizumab (D).

### SARS-CoV-2 RNAemia and N-antigenemia

A total of 340 plasma specimens from 73 patients (median, 4 samples/patient; range, 1-16) were available for analyses. SARS-CoV-2 RNAemia could be detected in 37 plasma specimens from 26 patients (35.6%). Median time to first detection of viral RNA in plasma was 10 days after symptoms onset (range, 3–32 days). The range of SARS-CoV-2 RNA loads measured in plasma was 1.69 to 5.27 log_10_ copies/ml. Median SARS-CoV-2 RNA load was significantly lower in plasma (3.03 log_10_ copies/ml) than in paired TA (Fig. 3A). A moderate yet significant correlation was found between SARS-CoV-2 RNA levels in TA and in paired plasma specimens (Fig. 3B). The dynamics of viral RNAemia are shown in Fig. 1B. SARS-CoV-2 RNAemia cleared at a faster rate than viral load in TA. In fact, viral RNAemia was not detected beyond day 32 after onset of symptoms. A similar conclusion was drawn taking only paired TA and plasma specimens into consideration for the analyses (Supplementary Fig. 3).

**Figure 3.**
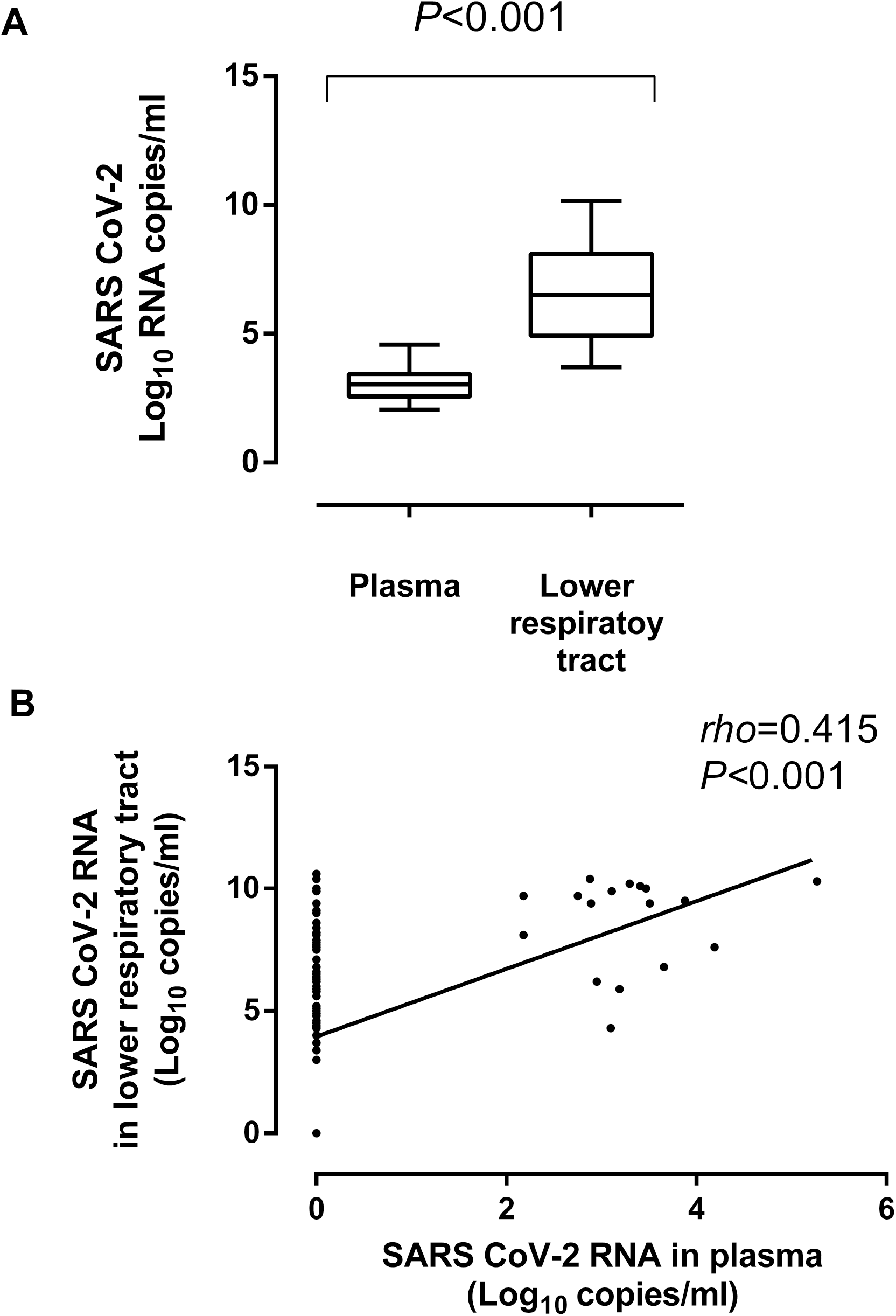
SARS-CoV-2 in the lower respiratory tract (tracheal aspirates) and plasma in critically ill patients undergoing invasive ventilation. Panel A shows SARS-CoV-2 RNA load in both compartments. Panel B shows the level of correlation between SARS-CoV-2 RNA load in tracheal aspirates and plasma specimens. *P* values and Spearman Rho value are shown.

SARS-CoV-2 RNA load in TA was significantly higher (*P*<0.001) in presence than absence of concomitant viral RNAemia (median, 9.5 log_10_ copies/ml; range, 4.3 to 10.4 log_10_ copies/ml vs. median, 6.2 log_10_ copies/ml; range, 3.0-10.6 log_10_ copies/ml).

SARS-CoV-2 N-antigenemia was detected in 43 samples from 30 patients (41.0%). Median time to first N-antigenemia detection was 9 days after onset of symptoms (range, 3–29 days). No patient had N-antigenemia beyond day 32 after symptoms onset. The level of agreement between the qualitative results returned by plasma RT-PCR and N-antigenemia assay was moderate (*k*=0.57; P=<0.0001). As shown in Fig. 4A, a trend towards higher SARS-CoV-2 RNA loads was seen in plasma specimens testing positive for N-antigenemia assay than in those yielding negative results (*P*=0.083). In turn, SARS-CoV-2 RNA load in TA was significantly higher (*P*<0.001) in the presence of concomitant N-antigenemia than in its absence (Fig. 4B).

**Figure 4.**
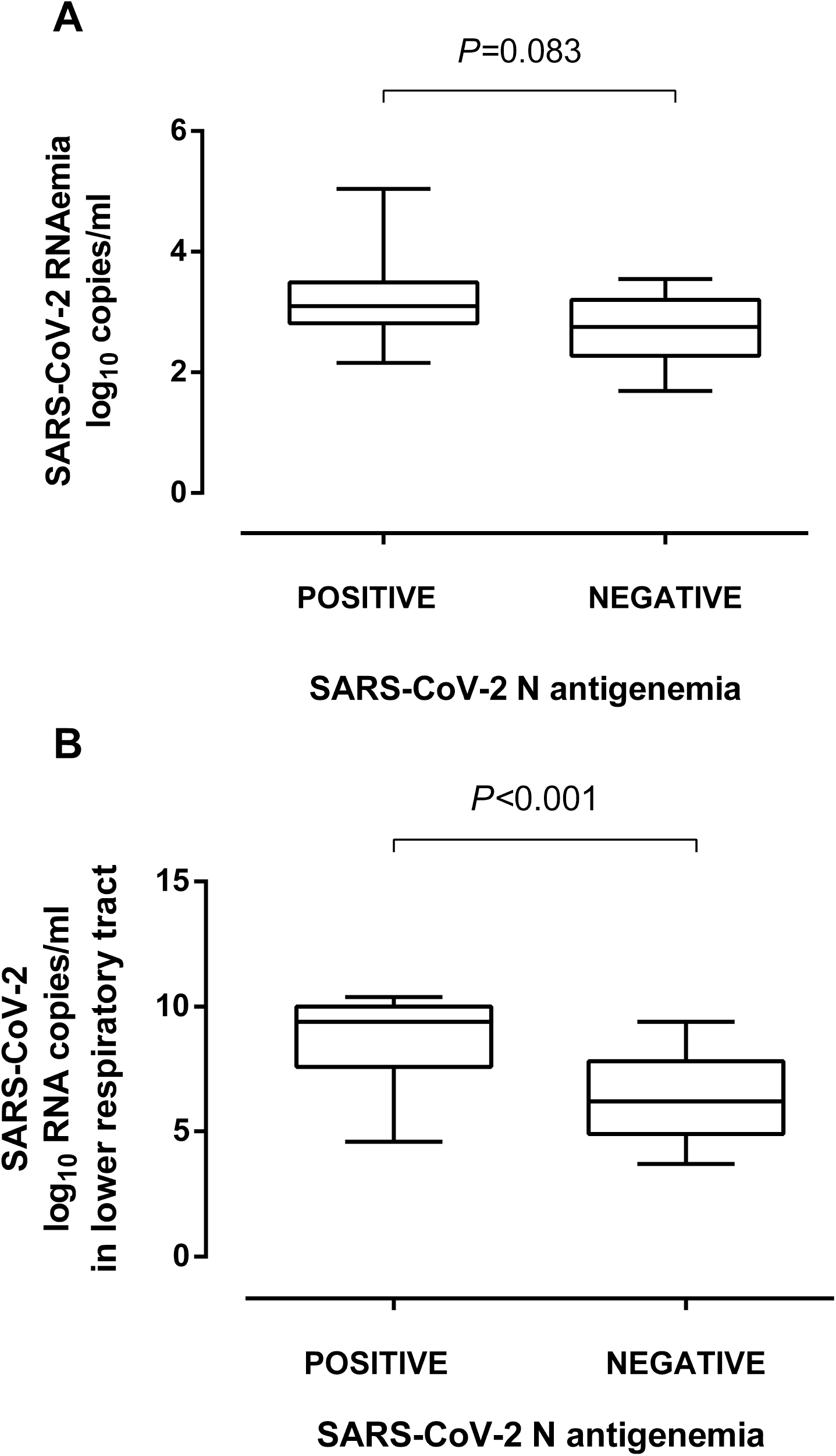
SARS-CoV-2 RNA load in plasma (A) and lower respiratory tract, i.e., tracheal aspirates (B) in critically ill patients undergoing invasive ventilation, in the presence or absence of SARS-CoV-2 N protein in paired plasma specimens. *P* values for comparisons are shown.

### Relationship between virological parameters, plasma levels of inflammatory or tissue-damage biomarkers and absolute lymphocyte counts

Plasma levels of ferritin and LDH, but not IL-6, CRP, or D-D, were significantly higher when SARS-CoV-2 RNA was detected in paired TA or plasma specimens (Table 2), yet SARS-CoV-2 RNA loads in TA and plasma correlated moderately with plasma levels of ferritin and LDH (Fig. 5A and 5B, respectively). The degree of correlation between SARS-CoV-2 RNA load and serum levels biomarkers was similar when all respiratory specimens (including NP) were considered for the analyses (not shown).

**Table 2.** Qualitative detection of SARS-CoV-2 RNA in the lower respiratory tract or plasma or SARS-CoV-2 N protein in plasma and blood levels of biomarkers of COVID-19 severity.

| Parameter<br>Median<br>(range) | Qualitative result of a given virological parameter | no. of paired specimens | SARS-CoV-2 RNA load in tracheal aspirates | *P* value | no. of paired specimens | SARS-CoV-2 RNAemia | *P* value | no. of paired specimens | SARS-CoV-2 N-antigenemia | *P* value |
| --- | --- | --- | --- | --- | --- | --- | --- | --- | --- | --- |
| IL-6 in pg/ml. | Pos | 23 | 111.4 (4-3,548) | 0.84 | 9 | 111.4 (10.8-1363.) | 0.92 | 14 | 105.8 (4-3,548) | 0.69 |
|  | Neg | 2 | 142 (22-262) |  | 40 | 141.8 (4-3,548) |  | 35 | 148.6 (4.7-3,437) |  |
| Ferritin in ng/ml. | Pos | 82 | 805.5 (69-6,440) | 0.01 | 31 | 1,176 (147-6,440) | <0.001 | 34 | 971.5 (147-6,440) | 0.03 |
|  | Neg | 34 | 421.5 (46-2,659) |  | 211 | 590 (42-5,847) |  | 208 | 599 (42-5,847) |  |
| D-D in ng/ml. | Pos | 101 | 1,730 (270-29,940) | 0.87 | 36 | 1,535 (320-9,170) | 0.17 | 41 | 1,350 (320-18,870) | 0.03 |
|  | Neg | 49 | 1,790 (270-16,160) |  | 274 | 1740 (270-60,000) |  | 269 | 1,760 (270-60,000) |  |
| LDH in UI/l. | Pos | 105 | 687 (93-2,132) | 0.001 | 36 | 765.5 (329-1,720) | 0.002 | 43 | 748 (214-1,720) | 0.002 |
|  | Neg | 51 | 520 (214-1,395) |  | 281 | 637 (58-2,132) |  | 274 | 632.5 (58-2,132) |  |
| CRP in mg/l. | Pos | 107 | 35 (1-746) | 0.62 | 36 | 47.95 (1.2-459) | 0.38 | 42 | 61.55 (1-459) | 0.01 |
|  | Neg | 54 | 32.85 (1-606.7) |  | 299 | 30.7 (1-746) |  | 293 | 29.9 (1-746) |  |
| Lymphocytes in cell/μl | Pos | 74 | 0.96 (0.02-3.73) | <0.001 | 26 | 0.72 (0.02-3.13) | <0.001 | 27 | 0.83 (0.02-1.65) | 0.001 |
|  | Neg | 42 | 1.40 (0.44-2.43) |  | 209 | 1.13 (0.17-3.73) |  | 208 | 1.12 (0.17-3.73) |  |

CRP, C-reactive protein; D-D, Dimer-D; IL-6, interleukin-6; LDH, lactose dehydrogenase.

**Figure 5.**
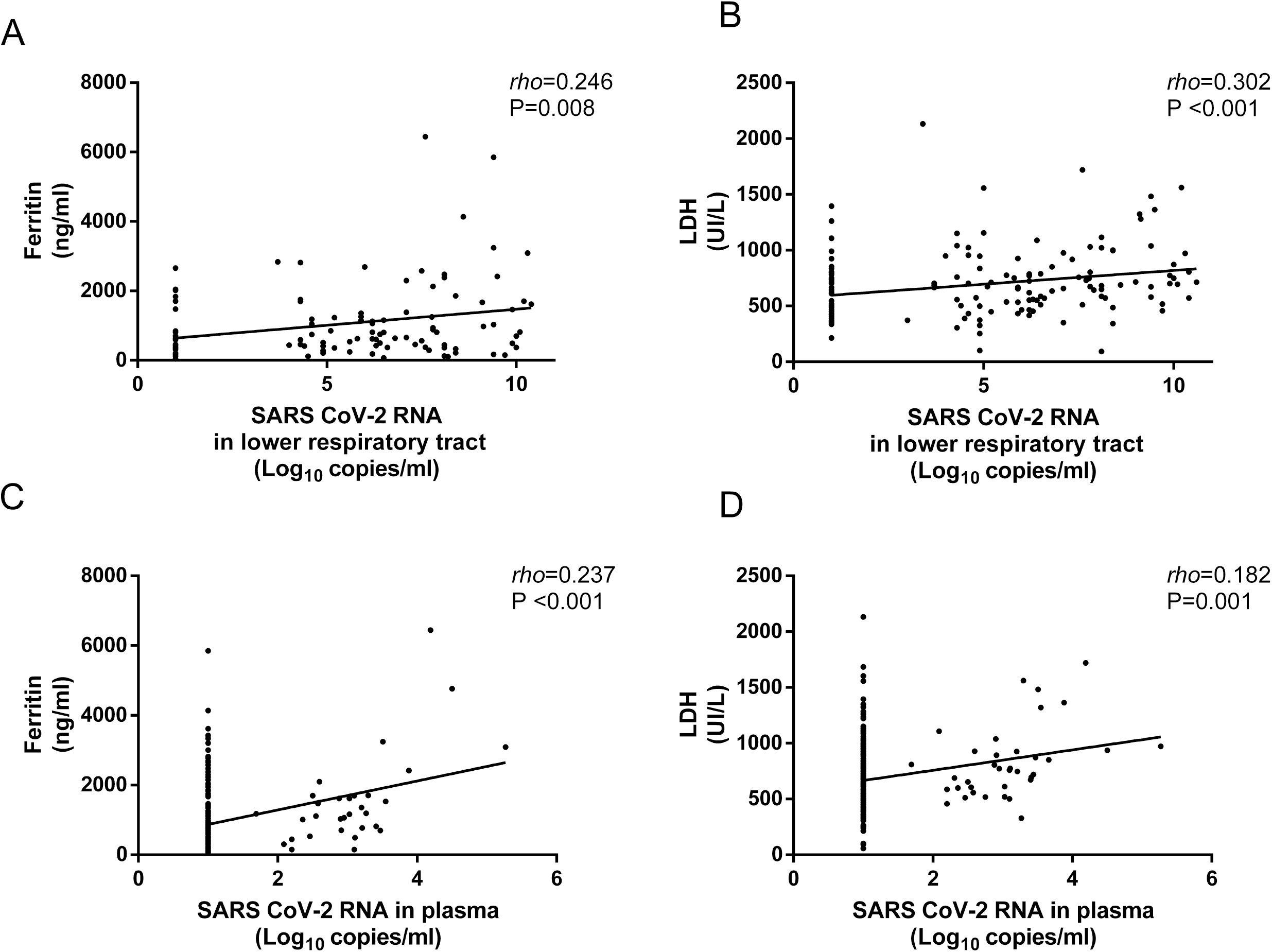
Correlation of serum levels of ferritin (A, C) and lactose dehydrogenase (LDH: B, D) with SARS-CoV-2 RNA load in paired lower respiratory tract specimens (tracheal aspirates) and plasma specimens from critically ill patients undergoing invasive ventilation. Spearman Rho and *P* values are shown.

Plasma levels of all biomarkers except IL-6 were significantly higher in specimens testing positive for SARS-CoV-2 N antigen than in those returning negative results (Table 2). Lymphocyte counts were significantly lower in the presence of SARS-CoV-2 RNA in TA and plasma or N antigen in plasma (Table 2). Nevertheless, the level of correlation (inverse) between SARS-CoV-2 RNA load in TA and plasma and lymphocyte counts was modest (Fig. 6).

**Figure 6.**
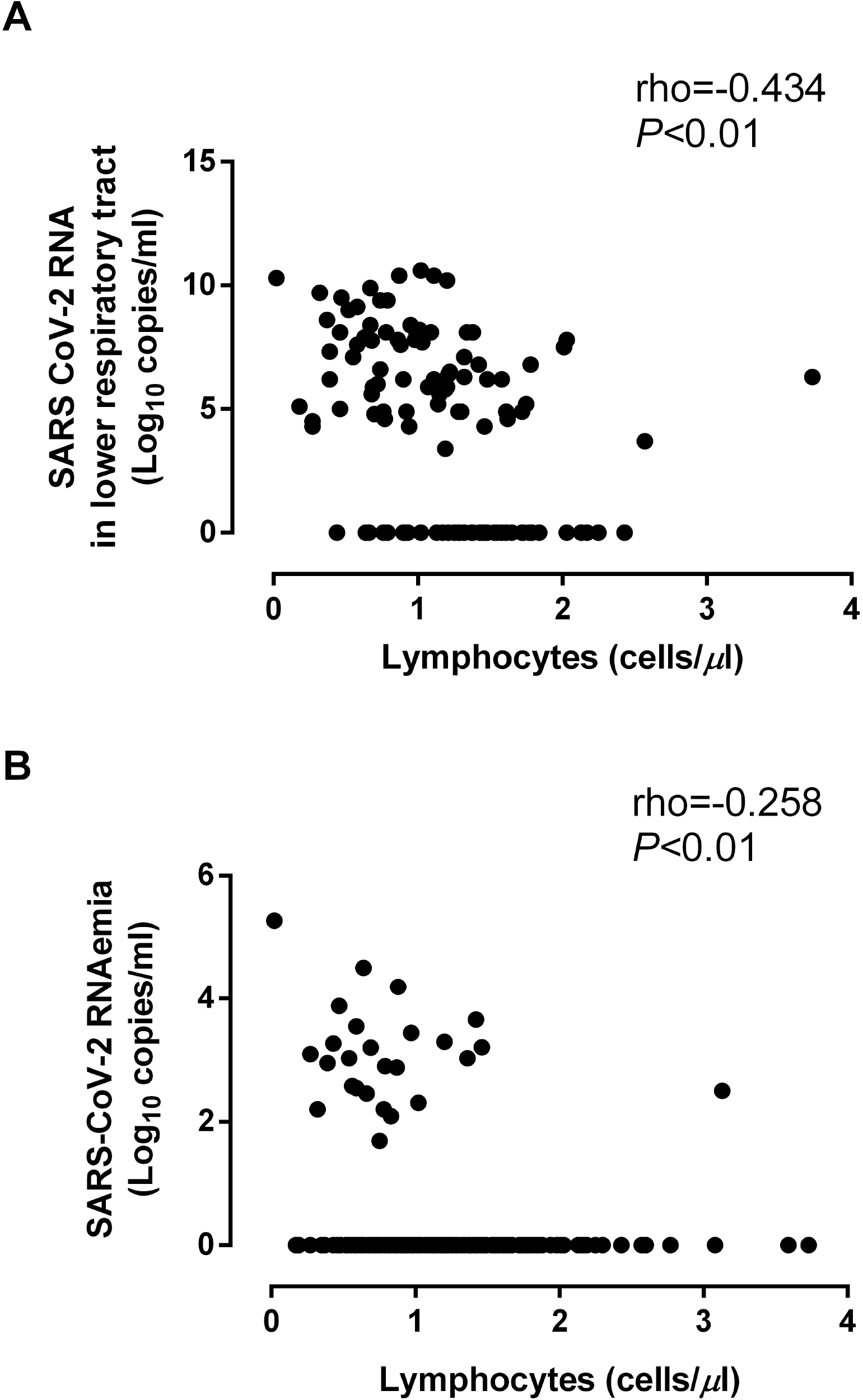
Correlation between SARS-CoV-2 RNA load in lower respiratory tract (upper panel) and plasma (lower panel) and absolute lymphocyte counts in paired specimens. Spearman Rho and *P* values are shown.

### Virological parameters and mortality

Dynamics of SARS-CoV-2 RNA load (initial, peak and trajectory) in TA following ICU admission were comparable across patients who either died or survived (Fig. 7 and Supplementary Fig. 4). Despite initial and peak SARS-CoV-2 RNA load in plasma being similar in surviving and deceased patients (*P*=0.11 and *P*=0.66, respectively; Supplementary Fig. 4), a trend towards a higher mortality rate (*P*=0.06) was noticed among patients exhibiting viral RNAemia (Table 3) in multivariate models adjusted for sex, age, comorbidities, APACHE II score and signs of organ failure. Occurrence of SARS-CoV-2 N-antigenemia was not associated with increased mortality (*P*=0.59).

**Table 3.** Risk factors for overall mortality among adult COVID-19 patients admitted to the intensive care unit.

| Variable | Univariate |  | Multivariate |  |
| --- | --- | --- | --- | --- |
|  | OR (CI, 95%) | *P* value | OR (95% CI) | *P* value |
| **Sex** |  |  |  |  |
| Males vs. females | 1.63 (0.56-4.66) | 0.36 | - |  |
| **Age** |  |  |  |  |
| ≥65 vs. < 65 years | 1.78 (0.69-4.58) | 0.23 | - |  |
| **Comorbidities (Yes vs. no)** | 1.44 (0.47-4.39) | 0.52 | - |  |
| Obesity | 0.78 (0.30-1.99) | 0.60 | - |  |
| Diabetes mellitus | 2.37 (0.80-6.99) | 0.11 | - |  |
| Hypertension | 1.55 (0.60-3.98) | 0.36 | - |  |
| Asthma/Chronic lung disease | 2.03 (0.56-7.42) | 0.28 |  |  |
| Chronic heart disease | 3.56(0.81-15.6) | 0.09 | 1.79 (0.33-9.68) | 0.49 |
| Vascular disease | 1.15 (0.24-5.58) | 0.85 |  |  |
| **Acute Physiology And Chronic Health Evaluation II** |  |  |  |  |
| Increase by one unit | 1.1 (0.98-1.22) | 0.09 | 1.03 (0.91-1.16) | 0.66 |
| <10 (ref) |  |  |  |  |
| 10-14 | 1.72 (0.43-6.91) | 0.44 |  |  |
| 15-29 | 1.94 (0.50-7.53) | 0.33 |  |  |
| ≤15 vs >15 | 2.17 (0.81-5.82) | 0.12 |  |  |
| **Organ failure** |  |  |  |  |
| Peak serum creatinine levels ≥1.3 vs. <1.3 | 3.87 (1.23-12.12) | 0.02 | 2.04 (0.54-7.67) | 0.29 |
| PiO<sub>2</sub>/FiO<sub>2</sub> <150 vs. ≥150 at admission | 0.583 (0.19-1.78) | 0.34 |  |  |
| Requirement of vasoactive medication within 24 h after ICU admission | 5.93 (2.12-16.6) | 0.001 | 6.34 (2.17-18.5) | 0.001 |
| Mechanical ventilation within 48 h of ICU admission | 2.34(0.67-8.15) | 0.18 |  |  |
| **Antiviral or anti-inflammatory treatments** |  |  |  |  |
| Tocilizumab (Yes vs. No) | 1.36 ( 0.52-3.59) | 0.52 |  |  |
| Remdesivir (Yes vs. No) | 0.71 (0.21-2.34) | 0.57 |  |  |
| **Virological parameters** |  |  |  |  |
| Increase by two units of SARS-CoV-2 RNA load (log<sub>10</sub> copies/mL) | 1.39 (0.78-2.48) | 0.26 |  |  |
| Initial SARS-CoV-2 RNA load in tracheal aspirates >7.1 vs. ≤7.1 log<sub>10</sub> copies/ml | 0.81 (0.29-2.24) | 0.68 |  |  |
| Initial SARS-CoV-2 RNA load in plasma (>3.0 vs. ≤3.0 log<sub>10</sub> copies/ml) | 1.68 (0.52-5.43) | 0.38 |  |  |
| SARS-CoV-2 RNA peak load in tracheal aspirates (≥7.5 vs. <7.5 log<sub>10</sub> copies/ml) | 0.63 (0.22-1.83) | 0.39 |  |  |
| SARS-CoV-2 RNA peak load in plasma (>5.6 vs. ≤5.6 log<sub>10</sub> copies/ml) | 0.53 (0.11-2.55) | 0.43 |  |  |
| SARS-CoV-2 N-antigenemia (Yes vs. No) | 1.29 (0.49-3.34) | 0.59 |  |  |
| SARS-CoV-2 RNAemia (Yes vs. No) | 2.49 (0.93-6.66) | 0.07 | 2.82 (0.94-8.47) | 0.06 |
| ICU, intensive care unit; OR, Odds ratio. |  |  |  |  |

**Figure 7.**
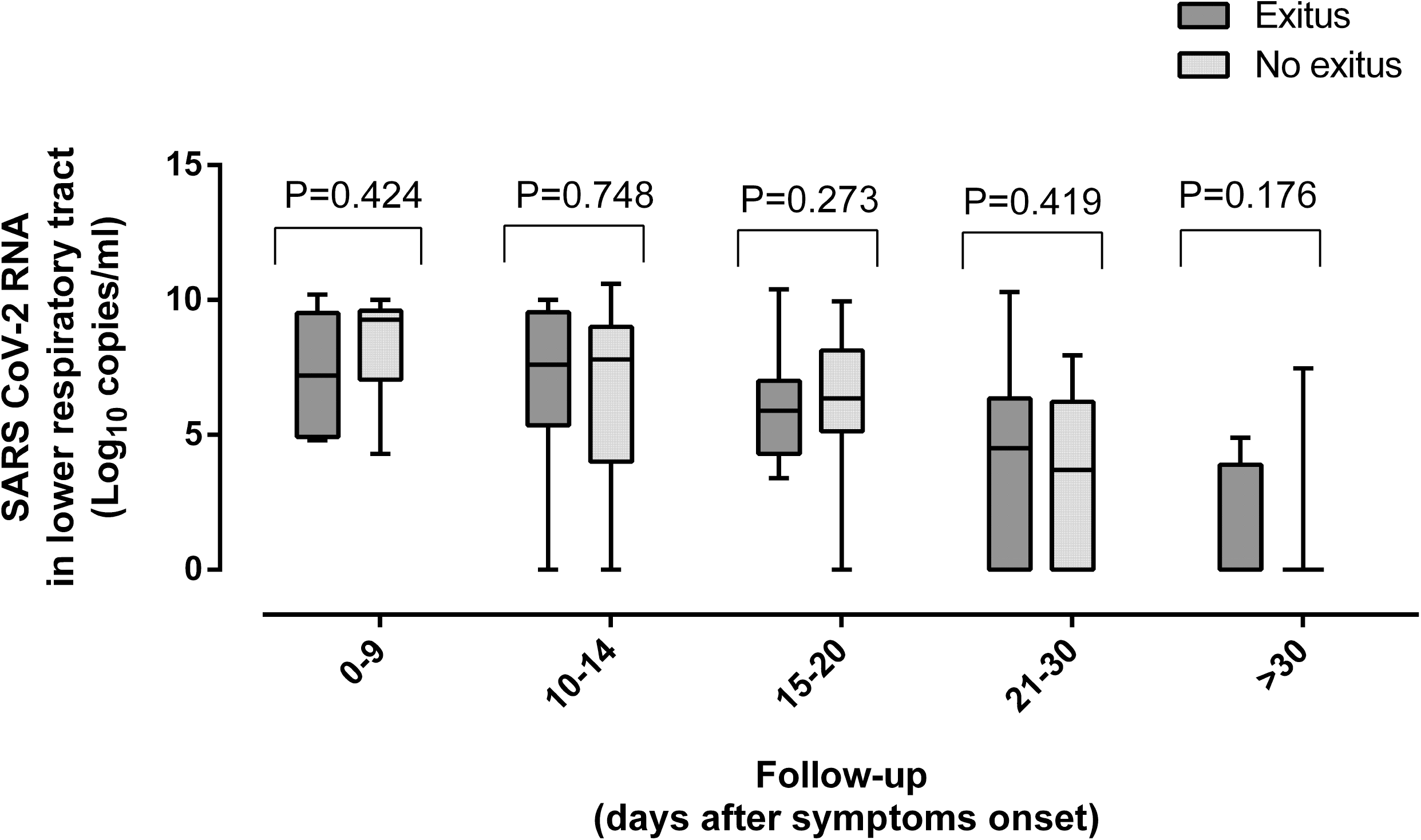
Kinetics of SARS-CoV-2 RNA load in the lower respiratory tract (tracheal aspirates) in critically ill patients undergoing invasive ventilation who either died or survived. *P* values for comparisons are shown.

## DISCUSSION

Here, we conducted a comprehensive comparative analysis of the kinetics of SARS-CoV-2 RNA load in the lower respiratory tract and blood compartment in a cohort of adult COVID-19 patients admitted to ICU, of whom almost 90% underwent invasive ventilation. The potential relationship between SARS-CoV-2 load in these compartments and systemic inflammation status, absolute lymphocyte counts and mortality was also investigated. In our view, the data presented herein could have relevant pathogenetic implications, and the following findings are underlined.

First, from a kinetics perspective, SARS-CoV-2 load in TA showed a descending trajectory from around two weeks after symptoms onset and was cleared in all survivors at seven weeks. Interestingly, neither remdesivir nor tocilizumab appeared to have a major impact on these kinetics. This is in line with previous studies showing that neither drug had a tangible effect on SARS-CoV-2 RNA load in the upper airways, even when administered early after symptoms onset [33,34].

SARS-CoV-2 RNAemia and N-antigenemia followed comparable time courses to viral load in TA, but they cleared more rapidly. Detection of both viral RNAemia and N-antigenemia was more likely in patients exhibiting higher SARS-CoV-2 loads in TA. Moreover, SARS-CoV-2 RNA load was substantially higher in TA than in paired plasma, and a moderate correlation was found between SARS-CoV-2 RNA load in TA and in paired plasma specimens. Collectively, these data suggested that LRT may be a substantial source of SARS-CoV-2 RNA or N protein circulating in the peripheral blood compartment. Given that detection of SARS-CoV-2 RNA in blood has not been associated with infectious virus [19], we speculate that free RNA and soluble N protein, probably leaking from the LRT (and perhaps from other tissue sources), are the main biological forms of SARS-CoV-2 present in plasma.

Second, SARS-CoV-2 RNAemia could be documented in one or more specimens from 35.6% of patients. This figure is substantially lower than was reported for ICU patients in other studies (77% in [14] and 88% in [18]). In these studies [14,18], however, a droplet-based digital PCR was used, which seemingly outperformed conventional RT-PCR assays such as the one used herein in terms of sensitivity. The above discrepancy could also be related to different timing of sample collection across studies, potentially important given the relatively rapid clearance of viral RNAemia documented in the current study.

Third, SARS-CoV-2 N-antigenemia was detected in 41.0% of patients overall by means of LFIC, with an analytical sensitivity of around 50 pg/ml: those exhibiting higher plasma viral RNA loads were more likely to test positive, as previously reported [28]. This concurs with the figure (49%) reported in a previous study [29] also using LFIC, but is much lower than the proportion found by Hingrat et al. [28] in a small subcohort of ICU patients (n=17), of whom most tested positive within the first two weeks after symptoms onset. Notably, an ultrasensitive immunoassay (limit of detection of 2.8 pg/ml) was employed in that study [28]. We did not measure serum antibodies against SARS-CoV-2, which may impact on the rate of N-antigenemia detection [29], and precludes certainty concerning the comparability of our data with those reported in the other studies [28,29]. Interestingly, in line with a previous report [28], N-antigenemia had a higher detection rate than viral RNAemia, which could be explained by the fact that N protein is likely less prone to degradation than RNA in cryopreserved-thawed specimens.

Fourth, we provide some evidence linking the presence of detectable SARS-CoV-2 RNA in TA and plasma, as well as N-antigenemia, with increased levels of inflammation or tissue damage biomarkers (ferritin and LDH, respectively), and decreased abolute lymphocyte counts, although the correlation between levels of these biomarkers and viral loads in TA and plasma was modest at best. Previous studies reported a significant association between SARS-CoV-2 RNAemia detection and blood levels of IL-6 [21] or several cytokines and chemokines, such as IL-6, IL-10, C-reactive protein, ferritin, D-D and LDH [14]. In these studies, however, a single time point specimen per patient collected at ICU admission was considered for the analyses, as opposed to the serial specimens used herein. Unfortunately, serum IL-6 levels were not available in a large percentage of patients of the current cohort.

Fifth, data from previous studies point to an association between protracted SARS-CoV-2 RNA clearance in LRT and/or simple presence of SARS-CoV-2 RNA in LRT and increased risk of mortality [9,10,34]. In these studies, a wide variety of LRT specimens were used, including sputa, TA and bronchoalveolar lavage. No data proving a dose-dependent relationship between SARS-CoV-2 RNA load in LRT and mortality were provided. Herein, we found no differences between surviving and deceased patients in initial SARS-CoV-2 RNA load after ICU admission, viral peak load or overall dynamics of SARS-CoV-2 RNA load in TA.

Sixth, both SARS-CoV-2 RNAemia and N-antigenemia have been independently associated with poor clinical outcome in mixed cohorts [see meta-analysis in 26] and in series including only ICU patients [14,29], in which patients who died displayed higher viral RNA loads in plasma collected at ICU admission than those who survived [14]. We found a trend towards an association between qualitative detection of SARS-CoV-2 RNA in plasma and mortality in multivariate logistic regression models, and failed to demonstrate higher initial or peak viral loads in deceased patients. We also found no evidence of an association between detection of N-antigenemia and mortality rate. Nevertheless, these data must be interpreted with caution due to the limited number of death events in our series.

The main limitations of the current study are the relatively small sample size, the non-negligible number of missing specimens, specially TA, compared with the planned sample collection schedule, and analytical methods which may have underestimated the viral RNAemia and N-antigenemia detection rate. Analysis of sequential specimens from patients could be considered a strength of the research.

The current study contributes to filling a knowledge gap on the comparative dynamics of SARS-CoV-2 in LRT and in the systemic compartment in ICU patients, and provides further insight into the pathogenesis of SARS-CoV-2 infection in this patient subset. In this respect, whether poor clinical outcomes in critically ill patients are directly linked to the extent of virus replication in the LRT, to a dysregulated pro-inflammatory state (cytokine storm) [4,5], or both, remains to be elucidated. In our view, our data fit better with a pathogenetic model, in which SARS-CoV-2 replication in the LRT or its presence in the blood compartment at a certain point over the course of ICU stay might not be a major driver of systemic inflammation, lymphopenia, lung dysfunction, multisystemic organ failure and death. This does not invalidate the importance of virus replication rate in the URT in the early stage after infection in determining the clinical course of COVID-19 [6-8]. Further studies are needed to resolve this issue.

## Data Availability

The data that support the findings of this study are available from the corresponding author, [author initials], upon reasonable request

## ACKNOWLEDGMENTS

No public or private funds were used for the current study. We are grateful to all personnel who work at Clinic University Hospital, in particular to those at Microbiology laboratory and the Intensive Care Unit for their commitment in the fight against COVID-19. Eliseo Albert holds a Juan Rodés Contract (JR20/00011) from the Health Institute Carlos III. Ignacio Torres holds a Río Hortega Contract (CM20/00090) the Health Institute Carlos III. We thank Siemens Healthineers (Erlangen, Germany) for providing reagents free of charge.

## AUTHOR CONTRIBUTIONS

BA, EA, IT, RG-R, RC, JC and JR: Methodology and validation of data. NC, JF and MLB: Medical care of ICU patients. DN: Conceptualization, supervision, writing the original draft. All authors reviewed the original draft.

## FIGURE LEGENDS

**Supplementary Figure 1.** Evaluation of the limit of detection of the CLINITEST^®^ Rapid COVID-19 Antigen Test (Siemens Healthineers, Erlangen, Germany) for detection of SARS-CoV-2 N protein in plasma specimens. A pre-pandemic plasma pool testing negative by RT-PCR was spiked with 10, 25, 50, 100 and 150 pg/ml of a recombinant N protein (MT-25C19NC, Certest Biotec S.L., Zaragoza, Spain).

**Supplementary Figure 2.** Representative example of a depleting experiment demonstrating the true nature of SARS-CoV-2 N protein detected in plasma specimens with N-antigenemia and testing negative for SARS-CoV-2 RNA by RT-PCR. A rabbit anti-N protein antibody (40143-R019; SinoBiological) was used for N protein depletion. An isotype-matched rabbit antibody was employed as a control.

**Supplementary Figure 3.** SARS-CoV-2 RNA load in plasma (A) and paired respiratory tract specimens (tracheal aspirates and nasopharyngeal exudates) (B) in critically ill patients undergoing invasive ventilation in the presence or absence of SARS-CoV-2 N protein in paired plasma specimens. *P* values for comparisons are shown.

**Supplementary Figure 4.** SARS-CoV-2 RNA peak load in plasma (upper panel) and tracheal aspirates (lower panel) in ICU patients who died or survived. *P* values for comparisons are shown.

## REFERENCES

1. Berlin DA, Gulick RM, Martinez FJ. Severe Covid-19. N Engl J Med. 2020;383:2451–2460.

2. Zheng S, Fan J, Yu F, et al. Viral load dynamics and disease severity in patients infected with SARS-CoV-2 in Zhejiang province, China, January-March 2020: retrospective cohort study. BMJ. 2020;369:m1443

3. Cevik M, Tate M, Lloyd O, Maraolo AE, Schafers J, Ho A. SARS-CoV-2, SARS-CoV, and MERS-CoV viral load dynamics, duration of viral shedding, and infectiousness: a systematic review and meta-analysis. Lancet Microbe. 2021;2:e13–e22.

4. Fajgenbaum DC, June CH. Cytokine Storm. N Engl J Med. 2020;383:2255–2273.

5. Leisman DE, Ronner L, Pinotti R, et al. Cytokine elevation in severe and critical COVID-19: a rapid systematic review, meta-analysis, and comparison with other inflammatory syndromes. Lancet Respir Med. 2020;8:1233–1244.

6. Alteri C, Cento V, Vecchi M, et al. Nasopharyngeal SARS-CoV-2 load at hospital admission as predictor of mortality. Clin Infect Dis. 2020 Jul 16:ciaa956.

7. Néant N, Lingas G, Le Hingrat Q, et al. Modeling SARS-CoV-2 viral kinetics and association with mortality in hospitalized patients from the French COVID cohort. Proc Natl Acad Sci U S A. 2021;118:e2017962118.

8. Pujadas E, Chaudhry F, McBride R, et al. SARS-CoV-2 viral load predicts COVID-19 mortality. Lancet Respir Med. 2020;8:e70.

9. Buetti N, Wicky PH, Le Hingrat Q, et al. SARS-CoV-2 detection in the lower respiratory tract of invasively ventilated ARDS patients. Crit Care. 2020; 24:610.

10. Huang Y, Chen S, Yang Z, et al. SARS-CoV-2 Viral Load in Clinical Samples from Critically Ill Patients. Am J Respir Crit Care Med. 2020;201:1435–1438.

11. Wang Y, Zhang L, Sang L, et al. Kinetics of viral load and antibody response in relation to COVID-19 severity. J Clin Invest. 2020;130:5235–5244.

12. Grant PR, Garson JA, Tedder RS, Chan PK, Tam JS, Sung JJ. Detection of SARS coronavirus in plasma by real-time RT-PCR. N Engl J Med. 2003;349:2468–2469.

13. Corman VM, Albarrak AM, Omrani AS, et al. Viral Shedding and Antibody Response in 37 Patients with Middle East Respiratory Syndrome Coronavirus Infection. Clin Infect Dis. 2016;62:477–483.

14. Bermejo□Martin JF, González□Rivera M, Almansa R, et al. Viral RNA load in plasma is associated with critical illness and a dysregulated host response in COVID□19. Crit Care. 2020; 24:691.

15. Berastegui□Cabrera J, Salto□Alejandre S, Valerio M, et al. SARS□CoV□2 RNAemia is associated with severe chronic underlying diseases but not with nasopharyngeal viral load. J Infect. 2020:S0163□4453:30719□2.

16. Hogan CA, Stevens BA, Sahoo MK, et al. High frequency of SARS□CoV□2 RNAemia and association with severe disease. Clin Infect Dis. 2020:ciaa1054

17. Xu D, Zhou F, Sun W, et al. Relationship between serum SARS□CoV□2 nucleic acid (RNAemia) and organ damage in COVID□19 patients: a cohort study. Clin Infect Dis. 2020:ciaa1085.

18. Veyer D, Kernéis S, Poulet G, et al. Highly sensitive quantification of plasma severe acute respiratory syndrome coronavirus 2 RNA sheds light on its potential clinical value. Clin Infect Dis. Clin Infect Dis. 2020:ciaa1196

19. Andersson MI, Arancibia□Carcamo CV, Auckland K, et al. SARS□CoV□2 RNA detected in blood products from patients with COVID□19 is not associated with infectious virus. Wellcome Open Res. 2020; 5: 181.

20. Prebensen C, Myhre PL, Jonassen C, et al. SARS□CoV□2 RNA in plasma is associated with ICU admission and mortality in patients hospitalized with COVID□19. Clin Infect Dis. 2020:ciaa1338.

21. Chen X, Zhao B, Qu Y, et al. Detectable serum SARS□CoV□2 viral load (RNAaemia) is closely correlated with drastically elevated interleukin 6 (IL□6) level in critically ill COVID□19 patients. Clin Infect Dis. 2020; 71: 1937□ 1942.

22. Hagman K, Hedenstierna M, Gille□Johnson P, et al. SARS□CoV□2 RNA in serum as predictor of severe outcome in COVID□19: a retrospective cohort study. Clin Infect Dis. 2020:ciaa1285.

23. Huang C, Wang Y, Li X, et al. Clinical features of patients infected with 2019 novel coronavirus in Wuhan, China. Lancet. 2020; 395: 497□ 506.

24. Chen W, Lan Y, Yuan X, et al. Detectable 2019□nCoV viral RNA in blood is a strong indicator for the further clinical severity. Emerg Microbes Infect. 2020; 9: 469□ 473.

25. Eberhardt KA, Meyer□Schwickerath C, Heger E, et al. RNAemia corresponds to disease severity and antibody response in hospitalized COVID□19 patients. Viruses. 2020; 12: 1045.

26. Tang K, Wu L, Luo Y, Gong B. Quantitative assessment of SARS-CoV-2 RNAemia and outcome in patients with coronavirus disease 2019. J Med Virol. 2021 Feb 16. doi:10.1002/jmv.26876

27. Ogata AF, Maley AM, Wu C, et al. Ultrasensitive serial profiling of SARS-CoV-2 antigens and antibodies in plasma to understand disease progression in COVID-19 patients with severe disease. Clin Chem. 2020;66:1562e72

28. Hingrat QL, Visseaux B, Laouenan C, et al. Detection of SARS-CoV-2 N-antigen in blood during acute COVID-19 provides a sensitive new marker and new testing alternatives. Clin Microbiol Infect. 2020:S1198-743X(20)30721-7.

29. Martin-Vicente M, Almansa R, Martínez I, et al. Absent or insufficient anti-SARS-CoV-2 S antibodies at ICU admission are associated to higher viral loads in plasma, antigenemia and mortality in COVID-19 patients. medRxiv 2021, https://doi.org/10.1101/2021.03.08.21253121

30. Mostafa HH, Hardick J, Morehead E, Miller JA, Gaydos CA, Manabe YC. Comparison of the analytical sensitivity of seven commonly used commercial SARS-CoV-2 automated molecular assays. J Clin Virol. 2020;130:104578.

31. Degli-Angeli E, Dragavon J, Huang ML, et al. Validation and verification of the Abbott RealTime SARS-CoV-2 assay analytical and clinical performance. J Clin Virol. 2020;129:104474.

32. Goldberg E, Ben Zvi H, Sheena L, et al. A real-life setting evaluation of the effect of remdesivir on viral load in COVID-19 patients admitted to a large tertiary center in Israel. Clin Microbiol Infect. 2021:S1198-743X(21)00113-0

33. Masiá M, Fernández-González M, Padilla S, et al. Impact of interleukin-6 blockade with tocilizumab on SARS-CoV-2 viral kinetics and antibody responses in patients with COVID-19: A prospective cohort study. EBioMedicine. 2020;60:10299

34. Bitker L, Dhelft F, Chauvelot L, et al. Protracted viral shedding and viral load are associated with ICU mortality in Covid-19 patients with acute respiratory failure. Ann Intensive Care. 2020;10:167.

