## Supplementary material for "SARS-CoV-2 RNA load in the lower respiratory tract, viral RNAemia and N-antigenemia in critically ill adult COVID-19 patients: relationship with biomarkers of disease severity": BEATRIZ OLEA 2021 SUPPLEMENTARY METHODS AND RESULTS.docx

Serum N protein depletion was achieved with an anti-N protein antibody produced in rabbit (40143-R019; SinoBiological). Two aliquots of 150 μL of COVID 19 positive serum were first incubated with 150 μL protein G agarose resin 4 rapid run (4RRPG; Agarose Bead Technologies) equilibrated in PBS for three hours at room temperature in batch mode for IgG depletion. Later, the mixtures were centrifuged for 10 min at 3,000 g at 4 ºC and the supernatants were collected. Then, 10 μg of anti-N protein antibody or rabbit IgG isotype (02-6102; Invitrogen) used as control were added to respective tubes and incubated overnight at 4 ºC in an orbital shaker. The samples were incubated again with 150 μL protein G agarose resin 4 rapid run equilibrated in PBS for three hours at room temperature in batch mode for specific IgG anti-N depletion and then centrifuged for 10 min at 3,000 g at 4 ºC and the supernatants were collected.

Serums were then analyzed for N protein presence with Clinitest® Rapid Covid-19 Antigen test (Siemens Healthineers), 150 μL of each serum were diluted 1:1 with extraction buffer, incubated for one minute at room temperature and 100 μL of each dilution were applied to the sample well of the lateral flow immunoassay.

After 15 minutes the results were analyzed in an Amersham Imager 680 UV (Ge Healthcare) with the software ImageQuant TL 8.2 (Ge Healthcare).

As shown in the Table below the depletion of N-protein from the serum with the anti-N protein antibody capture strategy reduced the test line intensity by 54 % compared with that obtained with the rabbit isotype control.

| **Sample** | **Line** | **Test line intensity %** |
| --- | --- | --- |
| α-N | Control | 93.24 |
| α-N | Test | 6.76 |
| Rabbit isotype | Control | 88.19 |
| Rabbit isotype | Test | 11.81 |

Table X. The bands of respective Clinitest® Rapid Covid-19 Antigen test were analyzed with the ImageQuant TL 8.2 software (Ge Healthcare). The volumes and percentage of each band are indicated in the table.
