## Supplementary material for "SARS-CoV-2 RNA load in the lower respiratory tract, viral RNAemia and N-antigenemia in critically ill adult COVID-19 patients: relationship with biomarkers of disease severity": BEATRIZ OLEA 2021 SUPPLEMENTARY TABLE 1.docx

| **Supplementary Table 1. Impact of treatment with remdesivir or tocilizumab on the dynamics of SARS-CoV-2 RNA load in the lower respiratory tract** | | | | | | |
| --- | --- | --- | --- | --- | --- | --- |
| Days (after symptoms onset) | **Treatment with remdesivir**  (SARS CoV-2 RNA  in tracheal aspirates in log_10_ copies/ml) | | | **Treatment with tocilizumab**  (SARS CoV-2 RNA  in tracheal aspirates in log_10_ copies/ml) | | |
|  | Yes | No | *P* value | Yes | No | *P* value |
| 0-9 | 9.45 (6.2-9.9) | 7.69 (4.3-10.20) | 0.46 | 9.4 (6.2-10.1) | 7.6 (4.3-10.2) | 0.20 |
| 10-14 | 7.6 (0-9.9) | 7.8 (0-10.6) | 0.80 | 8.1 (5-10) | 7.0 (0-10.6) | 0.13 |
| 15-20 | 6.2 (0-8.2) | 6.65 (3-10.4) | 0.33 | 6.8 (4.3-9.1) | 6.2 (0-10.4) | 0.21 |
| 21-30 | 4.4 (0-8.4) | 4.6 (0-10.3) | 0.89 | 4.9 (0-10.3) | 0 (0-8.4) | 0.06 |
| >30 | 0 (0-5.8) | 0 (0-8.1) | 0.72 | 1.85 (0-4.9) | 0 (0-8.1) | 0.12 |
