## Supplementary figures and images for "SARS-CoV-2 RNA load in the lower respiratory tract, viral RNAemia and N-antigenemia in critically ill adult COVID-19 patients: relationship with biomarkers of disease severity"

### BEATRIZ OLEA 2021 SUPPLEMENTARY FIGURE 1.TIF

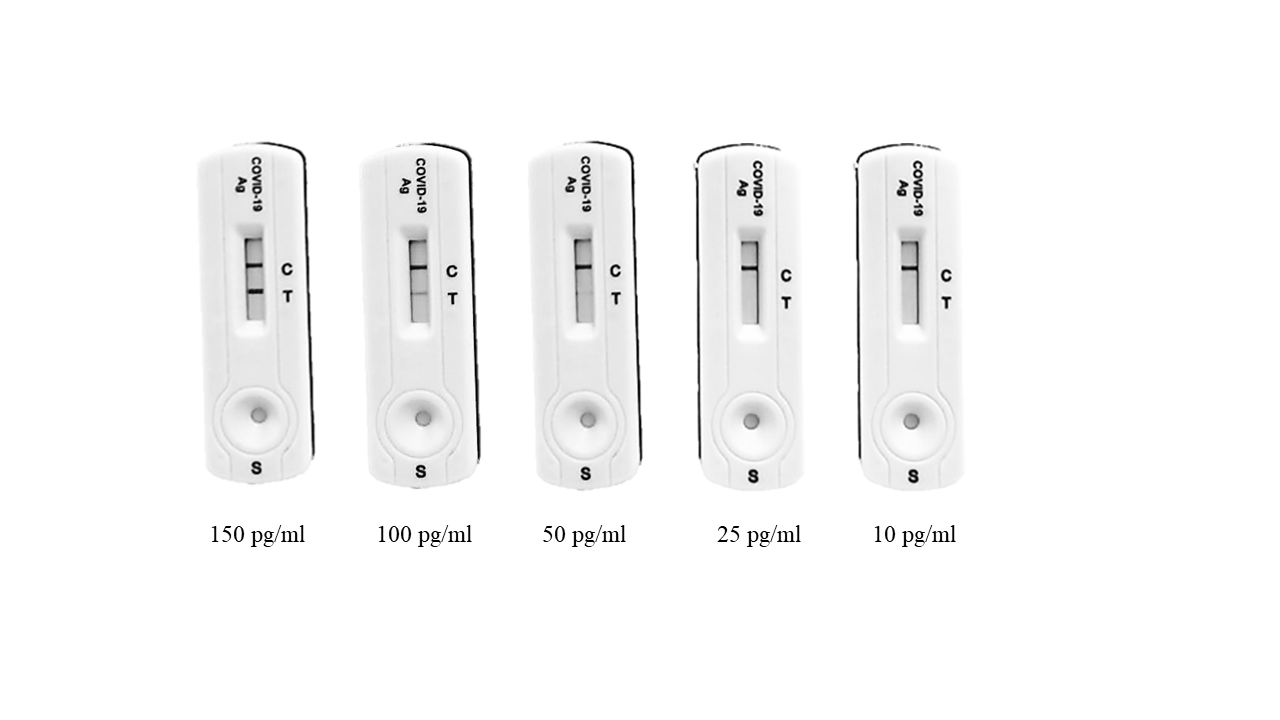

### BEATRIZ OLEA 2021 SUPPLEMENTARY FIGURE 2.TIF

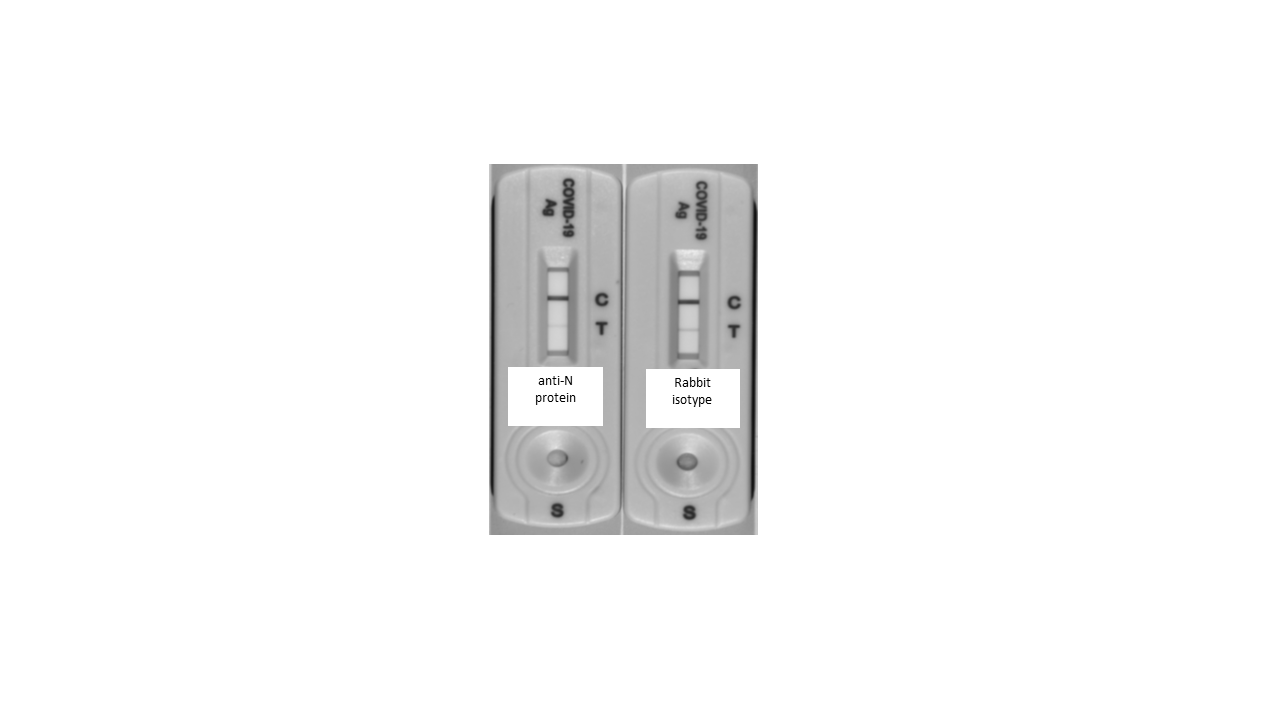

### BEATRIZ OLEA 2021 SUPPLEMENTARY FIGURE 3.tif

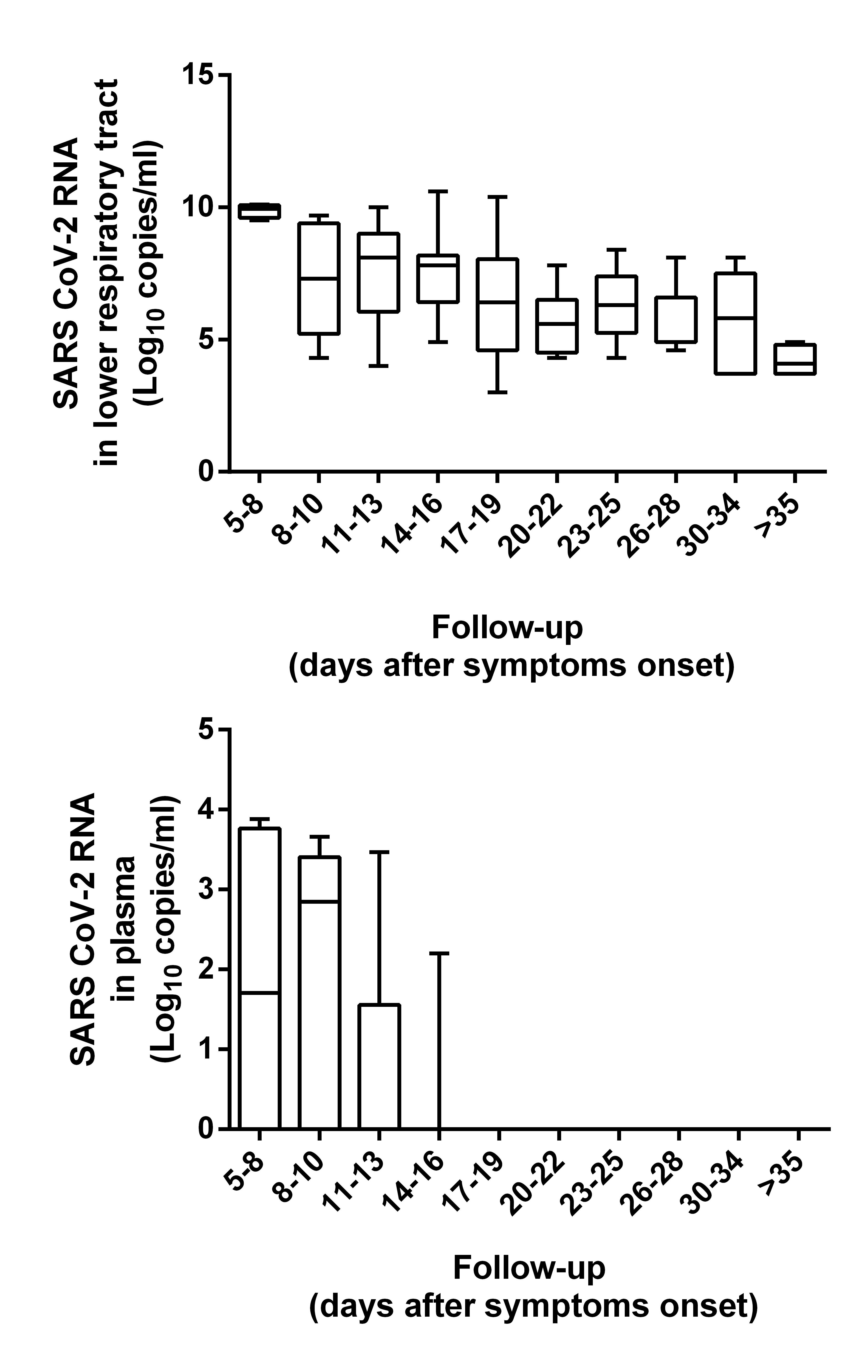

### BEATRIZ OLEA 2021 SUPPLEMENTARY FIGURE 4.tif

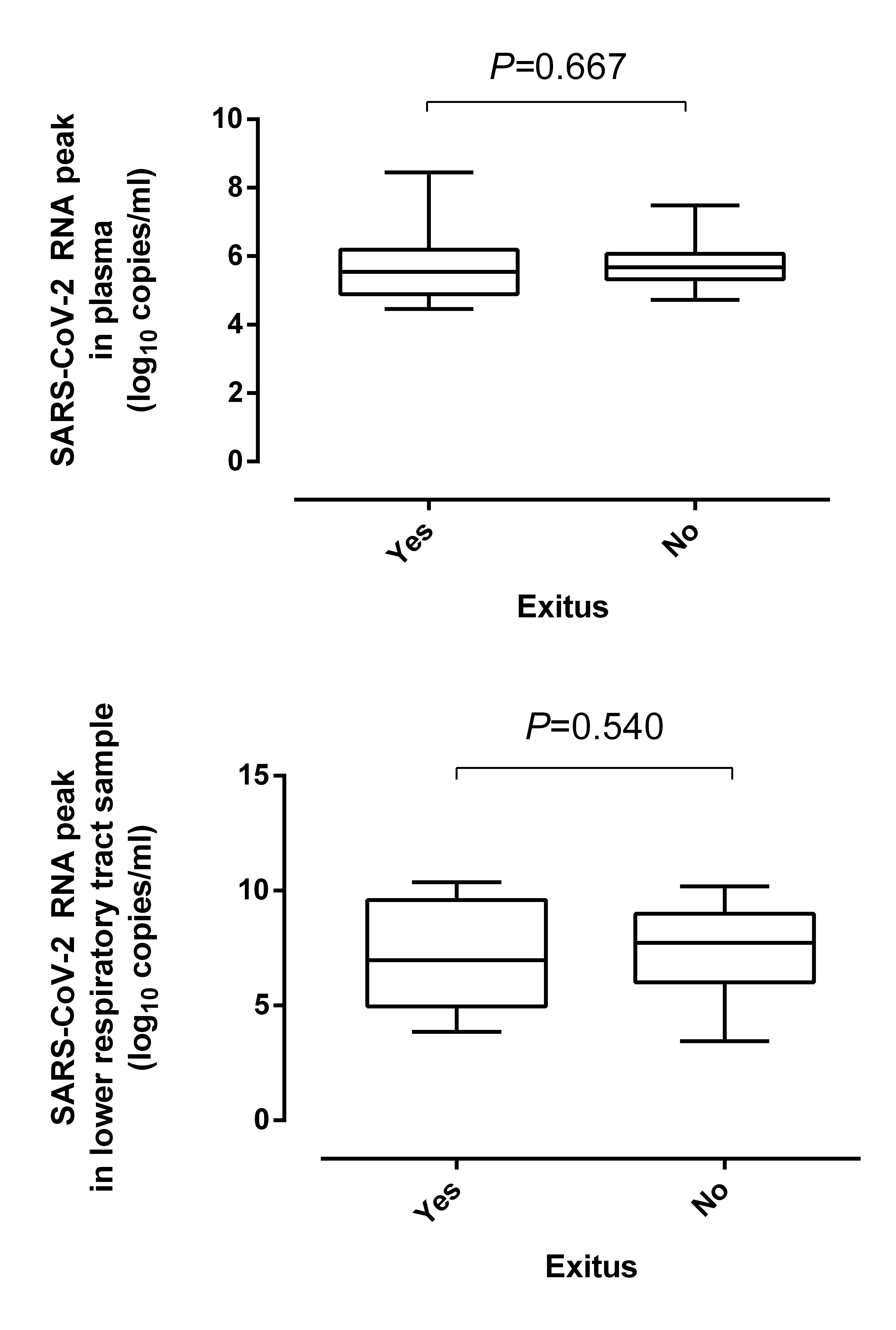
